# Spatiotemporal modelling and mapping of cervical cancer incidence among HIV positive women in South Africa: A nationwide study

**DOI:** 10.1101/2020.12.21.20248469

**Authors:** Dhokotera Tafadzwa, Riou Julien, Bartels Lina, Rohner Eliane, Chammartin Frederique, Johnson Leigh, Singh Elvira, Olago Victor, Muchengeti Mazvita, Egger Matthias, Bohlius Julia, Konstantinoudis Garyfallos

## Abstract

Disparities in invasive cervical cancer (ICC) incidence exist globally, particularly in HIV positive women who are at elevated risk compared to HIV negative women. We aimed to determine the spatial, temporal, and spatiotemporal incidence of ICC and the associated factors among HIV positive women in South Africa. We included ICC cases in women diagnosed with HIV from the South African HIV cancer match study during 2004-2014. We used the Thembisa model to estimate women diagnosed with HIV per municipality, age group and calendar year. We fitted Bayesian hierarchical models to estimate the spatiotemporal distribution of ICC incidence among women diagnosed with HIV. We also examined the association of deprivation, access to health (using the number of health facilities per municipality) and urbanicity with ICC incidence. We included 17,821 ICC cases and demonstrated a decreasing trend in ICC incidence, from 306 to 312 in 2004 and from 160 to 191 in 2014 per 100,000 person-years across all corrections. The spatial relative rate (RR) ranged from 0.27 to 4.43. In the model adjusting for covariates, the most affluent municipalities had a RR of 3.18 (95% Credible Interval 1.82, 5.57) compared to the least affluent ones, and municipalities with better access to health care had a RR of 1.52 (1.03, 2.27) compared to municipalities with worse access to health. More efforts should be made to ensure equitable access to health services, including mitigating physical barriers, such as transportation to health centres and strengthening of screening programmes.

**Novelty and Impact:** This is the first nationwide study in South Africa to evaluate spatial and spatiotemporal distribution of cervical cancer in women diagnosed with HIV. The results show an increased incidence of cervical cancer in affluent municipalities and in those with better access to health care. This is likely driven by better access to health care in more affluent areas. More efforts should be made to ensure equitable access to health services, including mitigating physical barriers.

## 1. Introduction

HIV positive women have an up to five times higher risk of developing cervical cancer (ICC) compared with HIV negative women.^1^ Persistent infection with high-risk variants of the human papillomavirus (HPV) is a necessary cause for ICC.^2^ In 2018, Southern Africa and Eastern Africa had the highest burden of cervical cancer worldwide with an incidence rate of over 40 cases per 100,000 population per year.^3^ The two regions also have a high burden of HIV: 54% of people living with HIV worldwide live in Southern and Eastern Africa.^4^

In 2019, the World Health Organization called for the elimination of cervical cancer as a public health threat.^5–7^ ICC can be prevented through HPV vaccination and early detection and treatment of pre-cancerous cervical lesions. When implemented effectively, these methods should result in decreased disease burden. However, globally there are disparities in ICC burden, even amongst HIV positive women. A global study demonstrated that HIV positive women in South Africa had a substantially higher incidence of ICC compared to the rest of the world, even after the introduction of antiretroviral therapy (ART).^8^ Poor access to screening, long waiting times for treatment of pre-cancerous cervical lesions as well as the socioeconomic inequities that exist in the country contribute to the high burden of ICC in South Africa.^9,10^

Spatiotemporal analysis of ICC incidence among HIV positive women can be used for disease surveillance over time and space, and to identify potential causes of the unequal distribution of ICC incidence. In this nationwide study in South Africa during 2004-2014, we examine the spatiotemporal incidence of ICC among women diagnosed with HIV. We perform three different corrections on the incidence rates to account for under-ascertainment, missing data and data errors. We assess whether urbanicity, deprivation and access to health services explained any geographical patterns.

## 2. Methods

### 2.1 Study Population

We retrieved data on women diagnosed with ICC with an HIV positive status in South Africa for the period 2004-2014 from the South African HIV cancer match (SAM) study. The SAM study aims to estimate cancer incidence in people living with HIV. It uses probabilistic record linkage methods to build a national cohort of people living with HIV from HIV-related laboratory tests stored at the National Health Laboratory Service (NHLS) and ICC cancer cases recorded in the National Cancer Registry (NCR).^11^ The HIV data includes laboratory HIV diagnostic tests like enzyme-linked immunosorbent assay (ELISA), qualitative and quantitative PCR as well as the Western Blot. In addition, the HIV data includes HIV laboratory monitoring tests such as CD4 cell counts and HIV RNA viral loads.^11^ The NCR is a nationwide, pathology-based cancer registry in South Africa with compulsory cancer reporting of cancer cases by both private and public laboratories since 2011.^12^ The NHLS is the largest diagnostic pathology service in South Africa, providing health services to about 80% of the population of South Africa (almost entirely users of public health facilities).^12^

We included ICC cases diagnosed within two years prior to a first HIV laboratory test or later, assuming that women would have been HIV-positive on average for at least two years prior to the test. The NHLS collects addresses of facilities where HIV tests are performed. We geocoded these addresses using Google Maps and linked them with corresponding municipalities. We assumed that the municipality of residence is the municipality where the women received the HIV-related test result.

We estimated the number of person-years of women diagnosed with HIV in each municipality using the Thembisa model (version 4.3).^13^ Thembisa is a mathematical model of the South African HIV epidemic, which provides provincial counts of cases diagnosed with HIV, stratified by age and sex. Since the Thembisa model does not provide estimates by municpality, we applied weights (by sex and municipality) using NHLS data on women diagnosed with HIV to obtain municipal HIV estimates (see Supporting Information Text S1.1 for the weight calculation and Figure S1 for the exclusion criteria of the NHLS).

### 2.2 Outcome

The NCR classifies cancer diagnoses according to the International Classification of Diseases for Oncology (ICD-O-3), excluding pre-cancerous lesions.^14^ We included cervical cancers coded C53.0, C53.1, C53.8 and C53.9.

### 2.3 Co-variates

We examined socioeconomic differentials across space using a multiple deprivation rank.^15^ The most deprived ward is given a rank of 1. To adjust for differences in access to health care, we considered the urbanicity of the municipalities (urban-rural) as retrieved from the National Department of Health data dictionary which was last updated in 2019 (Supporing Information Figure S2), and the number of health facilities per municipality 2004-2014 from NHLS.^16^ The health facility information in the NHLS dataset refers to primary and secondary care clinics and hospitals as well as regional and national hospitals from which the HIV test was requested. The spatial analysis was based on the addresses of these health facilities.

### 2.5 Statistical Methods

We used Bayesian hierarchical models to investigate the spatiotemporal distribution of ICC incidence among women diagnosed with HIV in South Africa. We first performed model selection without incorporating any covariates. Briefly, we used a re-parameterisation of the Besag-York-Mollié model to model the spatial autocorrelation.^17–19^ Random walk (RW) processes of order one were used to estimate the temporal random effect (on yearly resolution) and the random effects for the different age groups (15-19, 20-24, …, 75-79, >80 years). We fitted a series of models taking all possible combinations of the aforementioned random effects and also considered a type I interaction (interaction between an unstructured temporal and spatial random effects).^20^ To examine model performance and select the best performing model we derived the deviance information criterion (DIC), the Watanabe–Akaike information criterion (WAIC), and the mean logarithmic score (CPO).^21^ In a second step, we used the model that fitted best and adjusted for the selected covariates. We categorised the deprivation rank and number of health facilities per municipality in deciles to allow flexible fits. Supporting Information Text S1.2 provides further information about the model specification.

### 2.7 Incidence Corrections

We report four different estimates of the ICC incidence among women diagnosed with HIV. The first incidence estimates (‘no correction’) are from the best fitting model (without covariates).

The first correction (‘correction I’) accounts for the potential diagnosis of HIV in the private sector. NCR includes cases diagnosed in the private and public sector. The correction below assumes that if an NCR case is diagnosed in the private sector, then the women are likely to have received the HIV diagnosis and care in the private sector, too. Out of the total 57,161 ICC cases diagnosed during 2004-2014 in South Africa, 32,564 were not linked with an HIV case. However, out of the 32,564, only 5,077 (16%) were diagnosed in the private sector. We assumed that (57,161-32,564)/57,161=43% of these cases are HIV related. Thus, for correction I, we oversampled 5,077 *0.43=2,183 cases (on average 1,596 remained after the inclusion criteria) from the linked ICC cases that were treated in the private sector. To account for the sampling uncertainty, we fitted the model 100 times and averaged over the samples using Bayesian model averaging.^22^

For the second correction (‘correction II’), we used information about Kaposi sarcoma (KS). KS is a rare cancer in HIV-negative people but the most common cancer in people living with HIV.^23^ In SAM, 70% of the 23,046 KS cases could be linked probabilistically to an HIV test. Out of the KS cases not linked, 1,890 (8.2%) were diagnosed in the private sector. If we assume that all KS cases diagnosed from 2004-2014 occurred among people living with HIV, then the remaining 30%-8.2% = 21.8% missing HIV tests may be due to under-ascertainment of HIV, data errors or inaccuracies preventing linkage. To correct for the above, we calculated the proportion of unlinked KS cases by municipality excluding those treated in the private sector (because correction I accounts for that), and corrected the recorded numbers of ICC cases using the KS linked proportions (Supporting Information Text S1.3). The third correction combines corrections I and II (referred to as ‘full correction’).

We report median posterior and 95% credibility intervals (CrI) for the temporal ICC incidence rate and performed age standardisation based on the world population weights.^24^ We report median posterior spatial relative rates (RR; relative to the national average incidence rate over time), RR for the effect of the selected covariates (relative to the baseline) and posterior probabilities (that the RR is higher than 1). We used integrated nested Laplace approximation (INLA) for Bayesian inference.^25^ The data and the code used for the analysis are online available https://github.com/gkonstantinoudis/CervixHIVRSA.

## 3. Results

### 3.1 Study Population

We identified 57,161 ICC cases diagnosed from 2004-2014 in South Africa in the SAM study. We excluded 32,393 cases not linked with an HIV record, 1,607 with missing or imprecise geocodes, 3,666 with a ICC diagnosis two years or more before their first HIV related laboratory test and 1,674 cases residing in KwaZulu-Natal leaving us with 17,821 cases for the main analysis (Figure 1). We excluded the KwaZulu-Natal province because it started contributing data to NHLS only in 2010. With correction I, the total number of cases varied from 19,380 to 19,466. It was 21,446 for correction II and varied from 23,005 to 23,091 for the full correction. The total person-years (PY) at risk among women diagnosed with HIV during 2004-2014 were 24,059,679.

**Figure 1.**
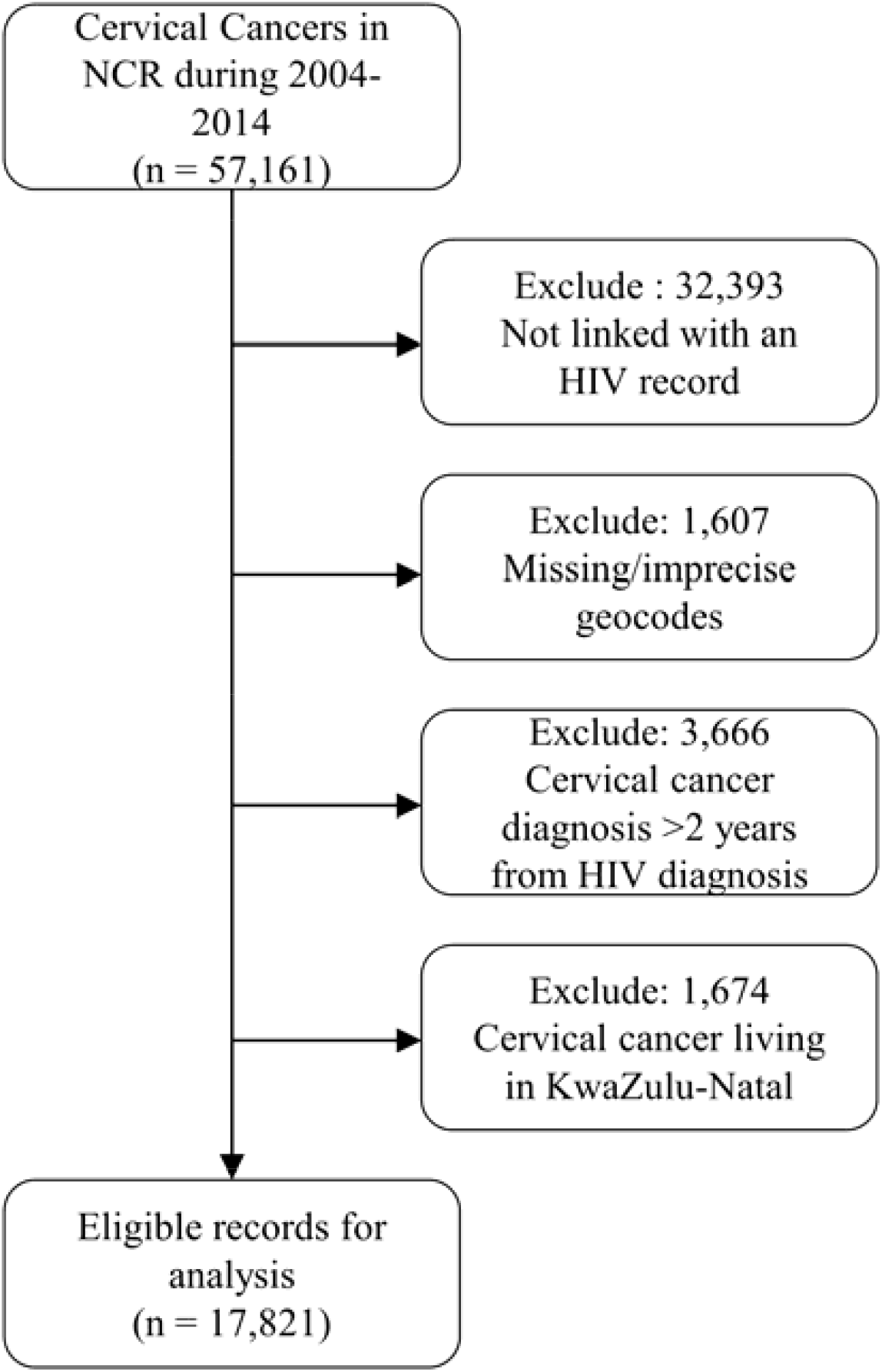
Flow of selection of eligible cervical cancer cases.

### 3.2 Model Selection

Based on the DIC, WAIC and CPO the model with the best fit was the one with random effects for age, time, space and a space-time interaction (Table S1 Supporting Information).

### 3.3 Co-variates

The spatial distribution of the deciles of deprivation rank and number of health facilities is shown in Figure 2. The spatial distribution of urbanicity is shown in the Supporting Information Figure S2. There were 23 urban and 190 rural municipalities in South Africa in 2016. Overall, we observe substantial spatial variation for all three covariates.

**Figure 2.**
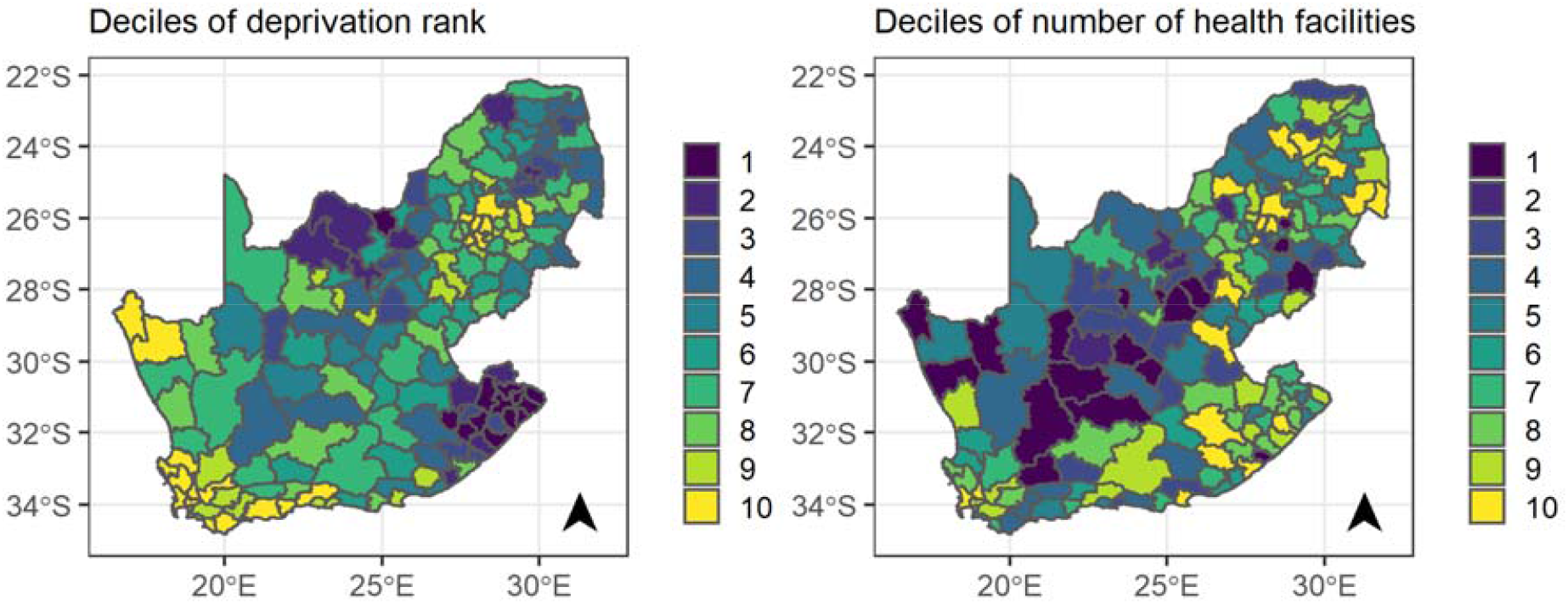
Maps of the deciles of the deprivation rank and number of health facilities per municipality.

### Overall Incidence

The median overall ICC incidence during 2004-2014 from the best fitting model was 294 (95% CrI: 174, 498) for no correction, 297 (95% CrI: 163, 573) for correction I, 310 (95% CrI: 170, 494) for correction II and 312 (95% CrI: 171, 510) for the full correction, per 100,000 PY, among women diagnosed with HIV (Table S2 Supporting Information).

### 3.3 Temporal Trends

Figure 3 shows that the annual median age-standardized ICC incidence rate per 100,000 PY of women diagnosed with HIV in South Africa decreased during 2004-2014. In 2004, the incidence rate varied from 306 (95% CrI: 169, 555) for no correction, to 306 (95% CrI: 163, 573) for correction I, and to 310 (95% CrI: 164, 589) for correction II and 312 (95% CrI: 160, 609) for the full correction, per 100,000 PY (Table S2 in Supporting Information). In contrast, in 2014 the incidence rate varied from 160 (95% CrI: 96, 265) for no correction, to 179 (95% CrI: 106, 303) for correction I, to 172 (95% CrI: 103, 290) for correction II and to 191 (95% CrI: 113, 326) for the full correction, per 100,000 PY (Table S2 in Supporting Information).

**Figure 3.**
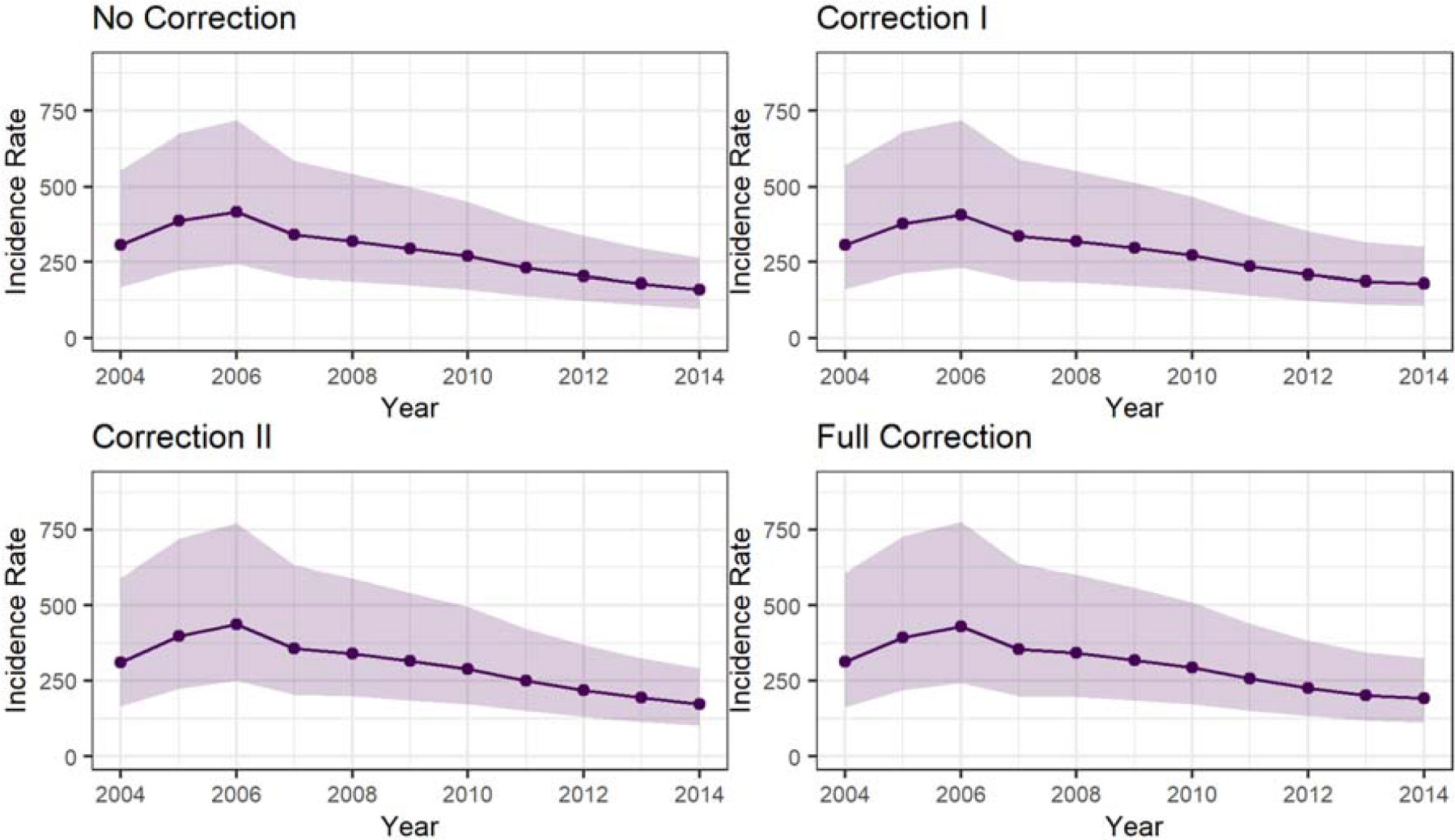
Yearly age-standardised incidence rate per 100,000 person years for women living with HIV in South Africa. Correction I refers to adjusting for those treated in the private sector, correction II for potential linkage errors and Full Correction is for both.

### 3.4 Spatial Patterns

Figure 4 shows the median posterior spatial RR and posterior probability of ICC across women diagnosed with HIV in the no correction model without covariates (top panels) and the no correction model adjusted for the selected covariates (bottom panels). For the model without covariates, the spatial RR ranged from 0.27 (95% CrI: 0.16, 0.42) to 4.43 (95% CrI: 3.71, 5.28). There was attenuation of associations when adjusting for covariates with the RR varying from 0.35 (95% CrI: 0.18, 0.64) to 4.10 (95% CrI: 2.90, 5.78). RR and posterior probabilities were larger in the province of the Northern Cape (north-western region of the map) and the Limpopo, Gauteng and Mpumalanga provinces (north-eastern part of the map) (Supporting Information Figure S3).

**Figure 4.**
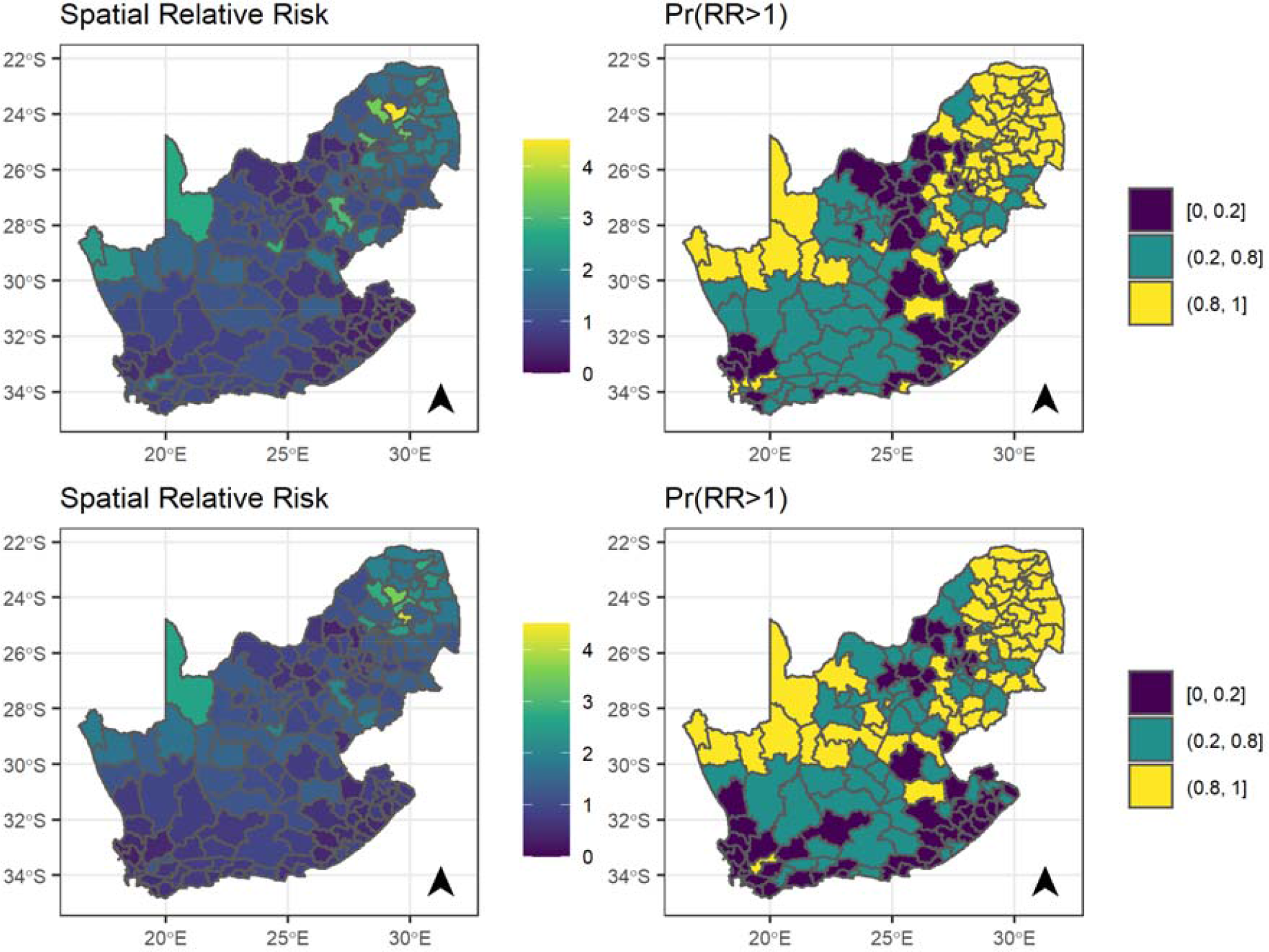
Median posterior of spatial relative risk (RR) and posterior probability that RR is larger than 1 of cervical cancer compared to the national average during 2004-2014 for the model without covariates (top panels) and the fully adjusted model (bottom panels). The axes denote the latitude and longitude, Pr: probability, RR: Relative Rate

### 3.5 Spatiotemporal patterns

The spatial differences have not attenuated over the years, and the patterns do not seem to be consistent in both ‘no correction’ models, with and without covariates (Supporting Information Figure S4-5).

### 3.6 Influence of covariates

We assessed the influence of covariates in the no correction model (Table 1). The RR comparing the least with the most deprived municipalities was 3.56 (95% CrI: 2.16-5.89) in the univariable model, and 3.18 (95% CrI: 1.82-5.57) in the multivariable model The RR in municipalities with a high number of health facilities was 1.83 (95% CrI: 1.27-2.64) in the univariable model, and 1.52 (95% CrI: 1.03-2.27) in the multivariable model, compared to municipalities with a low number of health facilities. There was little evidence of a difference in ICC incidence across urban and rural municipalities, both in the univariable (1.19, 95% CrI 0.94-1.5) and multivariable model (0.85, 95% CrI 0.65-1.12).

**Table 1.** Median posterior relative rate and 95% credibility intervals of the effect of the covariates.

| Co-variables |  | Univariate models |  |  | Multivariate model |  |  |
| --- | --- | --- | --- | --- | --- | --- | --- |
|  |  | Median | 95% CrI | Pr(RR>1) | Median | 95% CrI | Pr(RR>1) |
| Urbanicity | Rural | 1.00 | - | 1.00 | 1.00 | - | 1.00 |
|  | Urban | 1.19 | (0.94, 1.5) | 0.92 | 0.85 | (0.65, 1.12) | 0.12 |
| Deprivation rank | 1<sup>st</sup> Decile | 1.00 | - | 1.00 | 1.00 | - | 1.00 |
|  | 2<sup>nd</sup> Decile | 1.81 | (1.16, 2.82) | 1.00 | 1.77 | (1.13, 2.78) | 0.99 |
|  | 3<sup>rd</sup> Decile | 1.84 | (1.19, 2.86) | 1.00 | 1.68 | (1.07, 2.66) | 0.99 |
|  | 4<sup>th</sup> Decile | 2.14 | (1.35, 3.40) | 1.00 | 1.97 | (1.23, 3.16) | 1.00 |
|  | 5<sup>th</sup> Decile | 1.76 | (1.09, 2.83) | 0.99 | 1.83 | (1.10, 3.04) | 0.99 |
|  | 6<sup>th</sup> Decile | 2.22 | (1.41, 3.49) | 1.00 | 1.97 | (1.23, 3.17) | 1.00 |
|  | 7<sup>th</sup> Decile | 2.40 | (1.50, 3.85) | 1.00 | 2.33 | (1.43, 3.79) | 1.00 |
|  | 8<sup>th</sup> Decile | 2.82 | (1.78, 4.50) | 1.00 | 2.69 | (1.66, 4.35) | 1.00 |
|  | 9<sup>th</sup> Decile | 3.21 | (1.98, 5.21) | 1.00 | 2.88 | (1.75, 4.74) | 1.00 |
|  | 10<sup>th</sup> Decile | 3.56 | (2.16, 5.89) | 1.00 | 3.18 | (1.82, 5.57) | 1.00 |
| Number of health facilities | 1<sup>st</sup> Decile | 1.00 | - | 1.00 | 1.00 | - | 1.00 |
|  | 2<sup>nd</sup> Decile | 1.16 | (0.68, 1.98) | 0.71 | 1.14 | (0.67, 1.92) | 0.69 |
|  | 3<sup>rd</sup> Decile | 0.97 | (0.66, 1.42) | 0.43 | 0.90 | (0.62, 1.31) | 0.29 |
|  | 4<sup>th</sup> Decile | 1.14 | (0.77, 1.68) | 0.74 | 1.04 | (0.71, 1.55) | 0.59 |
|  | 5<sup>th</sup> Decile | 1.24 | (0.86, 1.81) | 0.87 | 1.06 | (0.73, 1.54) | 0.61 |
|  | 6<sup>th</sup> Decile | 1.17 | (0.79, 1.74) | 0.78 | 1.07 | (0.72, 1.60) | 0.64 |
|  | 7<sup>th</sup> Decile | 1.15 | (0.80, 1.67) | 0.77 | 1.05 | (0.72, 1.56) | 0.61 |
|  | 8<sup>th</sup> Decile | 1.31 | (0.91, 1.89) | 0.93 | 1.18 | (0.81, 1.71) | 0.81 |
|  | 9<sup>th</sup> Decile | 1.35 | (0.93, 1.97) | 0.95 | 1.24 | (0.85, 1.83) | 0.87 |
|  | 10<sup>th</sup> Decile | 1.83 | (1.27, 2.64) | 1.00 | 1.52 | (1.03, 2.27) | 0.98 |

CrI: Credibility intervals; Pr(RR>1): Posterior probability that the relative rate is larger than 1.

## Discussion

### 4.1 Main findings

In this study, we determined the spatial, temporal and spatiotemporal variation of ICC incidence in women diagnosed with HIV in South Africa. We observed an overall decreasing trend in ICC incidence in women diagnosed with HIV in South Africa during 2004-2014. We found a higher relative incidence in the Northern Cape and in the Limpopo, Gauteng and Mpumalanga provinces. Regarding the spatiotemporal patterns, the spatial differences did not decrease over time. Also, HIV positive women residing in the least deprived municipalities or in municipalities with larger numbers of health facilities were more likely to get diagnosed with ICC.

### 4.2 Strengths and limitations

To our knowledge, our study is the first nationwide study to describe the temporal, spatial and spatiotemporal variation in ICC incidence by municipality amongst women diagnosed with HIV in South Africa. Given that our data sources cover the majority of the South African population, and given the incidence adjustments we made, our data can be generalised to all women diagnosed with HIV in South Africa. The correction factors reduced the selection bias introduced through our data sources and the record linkage process. However, our study also has several limitations. In our study, the population under observation was specific to a sub-population of women who knew their HIV status and thus had contact with health care facilities. This contact increases the probability of being referred for ICC screening and treatment when compared to women who are unaware of their HIV status. As a result, the results cannot be generalised to women who have not been diagnosed with HIV. According to a survey by the Human Science Research Council, 47.5% of women aged 15 years and older were unaware of their HIV positive status in 2017.^26^ Although we introduced several incidence corrections, women without health care access are missing from our study. The ecological nature of our analysis did not allow us to adjust for CD4 counts that might influence ICC incidence. We could not adjust for screening coverage due to the absence of nationwide screening data for women living with HIV.

### 4.3 Discussion in the context of previous studies

The high incidence observed across all corrections is consistent with other studies on ICC incidence in women living with HIV in South Africa. Two hospital-based cohort studies determined crude incidence rates that ranged from 447-506 ICC cases/100 000 person-years.^8,27^ These results reflect the high ICC burden in HIV positive women in South Africa compared to the overall crude incidence rate of ICC in the general South African population. The latter experienced an incidence of 25.57 ICC cases/ 100 000 of the population as of 2016.^28^ Other HIV and cancer registry linkage studies in Africa have shown variable crude incidence rates in women living with HIV. In Nigeria, investigators observed an incidence of 7.8/ 100 000 person-years^29^ whilst in Uganda, the incidence reported was 70/ 100 000 person-years, with the incidence being highest in the 35-44 age group at 200/ 100 000 person-years^30^. These differences in ICC incidence might potentially reflect differences in access to ICC screening and diagnosis in the different countries

We observed a decreasing trend in ICC incidence after an initial increase between 2004 and 2006, similar to the decline that was observed in the global North after the introduction of ART.^31,32^ However, in other African settings, the ICC incidence trends either remained unchanged or increased even after the introduction of ART.^30,33^ The decline in incidence in our study might reflect the HIV testing patterns in South Africa across the study period. Initially, fewer people were tested for HIV,^34^ and those tested had an increased burden of disease and lower CD4 cell counts.^35^ Increased ICC incidence has been associated with very low CD4 cell counts compared to higher CD4 cell counts. With time, as HIV testing expanded, more women had access to timely HIV diagnosis and treatment before advanced disease, thus reducing the likelihood of ICC. The timely access might have resulted in the subsequent decline in ICC incidence from 2006 amongst women diagnosed with HIV. ART reduces the prevalence of high-risk HPV infections and cervical pre-cancerous lesions. Still, the effect is reduced in individuals initiating ART at an advanced HIV stage and in women with advanced pre-cancerous lesions.^36–38^ The expansion of the national ART programme, which started in 2004, ensured that more people had access to HIV treatment and subsequently improved CD4 cell counts. This expansion could also explain the subsequent decline in incidence from 2006-2014 as women had a lower burden of disease and better access to ART. However, in some studies this has not been supported. In Botswana, no significant change in ICC incidence was observed after ART expansion.^33^

Spatial analyses have been used in ICC studies to determine disparities in incidence and mortality as well as the factors associated with the unequal distribution of ICC burden.^27,39–42^ However, all these studies focused on the general population regardless of HIV status. Previous studies examining the spatial trends in the USA, Thailand and Cuba have reported variations in ICC incidence driven by a variety of factors.^39,43,44^ In Thailand, Zhao et al. observed differences in ICC incidence in districts in the Songhkla province driven by sexual behaviour and migration patterns.^44^ A similar study in Cuba showed that differences in screening programme coverage did not account for the spatial differences in ICC incidence, which were better explained by differences in lifestyle and socioeconomic status.^40^ In the USA, the disparities were attributed to ethnicity, socioeconomic status and screening coverage.^39^ Low socioeconomic status of areas and low screening rates were associated with an increased ICC incidence. Another spatiotemporal study in Texas, USA demonstrated that poor education, African and Hispanic ethnicities and low socioeconomic status was associated with a high ICC incidence.^45^

In our study, more affluent areas had a higher incidence of ICC. People living in the least deprived municipalities were about three times more likely to be diagnosed with ICC. This points towards disparities in ICC, driven by unequal access to services due to socioeconomic circumstances. In Uganda, a spatial analysis of ICC in women, regardless of HIV status, showed that areas of low socioeconomic status had decreased access to care, including ICC screening.^41^ This can lead to a spurious negative association between ICC incidence and socioeconomic status, as areas of low socioeconomic status would appear to have a lower burden of disease when the ICC cases are not being diagnosed. Some districts in South Africa have pronounced shortages of health care staff, with multiple vacant posts and ill-equipped facilities,^46^ which affects access to care. In addition, utilisation of public health sector facilities in South Africa has been shown to be higher among people from least deprived areas, with individuals from deprived areas having limited access to public health sector services.^47^

We observed a relationship between the number of facilities per municipality and ICC incidence in women diagnosed with HIV: the greater the number of health facilities, the higher the ICC incidence. This points to the effect of inequalities in access to care on ICC incidence. In the United States, in urban areas, a larger distance to the health facility was a barrier to accessing cervical cancer care and treatment.^48^ However, the same was not observed in rural areas, where long distances were associated with better access, with the suggested reason being the familiarity and tolerance to long-distance travel for health services in rural areas.^48^ A qualitative study in Botswana determined thematic issues that affected access to ICC treatment and care. Among those were contextual issues like distance from facility, other commitments and transportation issues as well as health system issues such as long waiting times for appointments with health care workers and for test results.^49^ In South Africa, it is estimated that for every additional health care professional per year, the likelihood of cervical cancer screening improved by 1%.^50^ In our study, we could not assess any of the aforenmentioned different metrics pertaining to access. We should also note that we assumed that the municipality of the health facility is the same as the municipality of residence. This assumption may be problematic as people can move to other provinces and municipalities for healthcare. For instance, municipalities with more facilities might have better accessibility and shorter waiting times and hence people might choose to go there instead. Thus, we cannot rule out that the observed effect can be driven by this assumption.

Deprivation, access to health and urbanicity did not explain all of the observed spatial variation. The remaining spatial trends can point towards geographical health inequalities, potentially generated by different screening programmes and municipality level health policies. Differences in HPV prevalence might also influence the ICC incidence patterns. There was no readily available nationwide screening coverage data or ethnicity data at the municipality level for women diagnosed with HIV at the time of analysis.

## 4.4 Conclusion

In conclusion, although the burden of ICC among women diagnosed with HIV has decreased over time, there was strong spatial variation. Individuals from less deprived areas and areas with better access to health facilities are more likely to be diagnosed with ICC compared to those from most deprived areas or areas with worse access to health facilities. The geographical discrepancies persisted after adjusting for the aforementioned factors, which could reflect spatial differences in screening policies across women living with HIV. More efforts should be made to ensure equitable access to health services, including mitigating physical barriers, such as transportation to health centres, strengthening of screening programmes, and nationwide HPV vaccination implementation in populations living with HIV. Special attention should be paid to women living in low socioeconomic areas.

## Data Availability

The data will be made available as aggregated data. Additional data can be granted upon reasonable request

https://github.com/gkonstantinoudis/CervixHIVRSA

## Authors’ contributions

Conceptualisation: GK, ME, ES and JB; Data Acquisition: MM, ES and LJ (Thembisa estimates). Methodology: LB and VO were involved in data cleaning and conducted record linkages; Formal analysis and validation: GK; Writing-original draft: GK and TD. All authors contributed towards data interpretation and critical comments on the first and/or subsequent drafts. All authors read and approved the final manuscript.

## Ethical approval

Permission to use the routinely collected NHLS and NCR data was sought from the relevant authorities. Ethical approval to conduct the study was granted by the University of the Witwatersrand Human Research Ethics Committee [Ethics certificate numbers (SAM: M160944)].

## Acknowledgements

The authors would like to thank all funders, the National Health Laboratory Service (NHLS), the NHLS’s Corporate Data Warehouse and the National Cancer Registry.

## References

1 Ghebre RG, Grover S, Xu MJ, Chuang LT, Simonds H. Cervical cancer control in HIV-infected women: Past, present and future. Gynecol. Oncol. Reports. 2017; 21: 101–8.

2 Bouvard V, Baan R, Straif K, et al. A review of human carcinogens—Part B: biological agents. Lancet Oncol 2009; 10: 321–2.

3 Bray F, Ferlay J, Soerjomataram I, Siegel RL, Torre LA, Jemal A. Global cancer statistics 2018: GLOBOCAN estimates of incidence and mortality worldwide for 36 cancers in 185 countries. CA Cancer J Clin 2018; 68: 394–424.

4 UNAIDS. 2018 Global HIV Statistics. Unaids 2019; : 1–6.

5 Brisson M, Drolet M. Global elimination of cervical cancer as a public health problem. Lancet Oncol. 2019; 20: 319–21.

6 Hall MT, Simms KT, Lew J-B, et al. The projected timeframe until cervical cancer elimination in Australia: a modelling study. Lancet Public Heal 2018; 0. DOI:10.1016/S2468-2667(18)30183-X.

7 Simms KT, Steinberg J, Caruana M, et al. Impact of scaled up human papillomavirus vaccination and cervical screening and the potential for global elimination of cervical cancer in 181 countries, 2020–99: a modelling study. Lancet Oncol 2019; 20: 394–407.

8 Rohner E, Sengayi M, Goeieman B, et al. Cervical cancer risk and impact of Pap-based screening in HIV-positive women on antiretroviral therapy in Johannesburg, South Africa. Int J Cancer 2017; 141: 488–96.

9 Olorunfemi G, Ndlovu N, Masukume G, Chikandiwa A, Pisa PT, Singh E. Temporal trends in the epidemiology of cervical cancer in South Africa (1994–2012). Int J Cancer 2018; 143: 2238–49.

10 Jordaan S, Michelow P, Simoens C, Bogers J. Challenges and Progress of Policies on Cervical Cancer in South Africa. Heal Care Curr Rev 2017; 05. DOI:10.4172/2375-4273.1000188.

11 Muchengeti M, Bartels L, Olago V, et al. Cohort Profile: The South African HIV Cancer Match Study (SAM). DOI:10.31219/OSF.IO/W52SB.

12 Singh E, Ruff P, Babb C, et al. Establishment of a cancer surveillance programme: The South African experience. Lancet Oncol. 2015; 16: e414–21.

13 Johnson LF, Dorrington RE, Moolla H. Progress towards the 2020 targets for HIV diagnosis and antiretroviral treatment in South Africa. South Afr J HIV Med 2017; 18: 1–8.

14 Fritz A, Percy C, Jack A, et al., editors. International Classification of Diseases for Oncology: Third Edition, Third. 2013 https://apps.who.int/iris/handle/10665/42344.

15 Noble M, Wright G. Using Indicators of Multiple Deprivation to Demonstrate the Spatial Legacy of Apartheid in South Africa. Soc Indic Res 2013; 112: 187–201.

16 National Department of Health. National Department of Health Data Dictionary. 2019. https://dd.dhmis.org/# (accessed July 6, 2020).

17 Besag J, York J, Mollie A. Bayesian image restoration with two applications in spatial statistics. Ann Inst Stat Math 1991; 43: 1–20.

18 Simpson D, Rue H, Riebler A, Martins TG, Sorbye SH. Penalising Model Component Complexity: A Principled, Practical Approach to Constructing Priors. Stat Sci 2017; 32: 1–28.

19 Konstantinoudis G, Schuhmacher D, Rue H, Spycher BD. Discrete versus continuous domain models for disease mapping. Spat Spatiotemporal Epidemiol 2020; 32: 100319.

20 Knorr-Held L. Bayesian modelling of inseparable space-time variation in disease risk. Stat Med 2000; 19: 2555–67.

21 Gelman A, Hwang J, Vehtari A. Understanding predictive information criteria for Bayesian models. Stat Comput 2014; 24: 997–1016.

22 Gómez-Rubio V, Bivand RS, Rue H. Bayesian model averaging with the integrated nested Laplace approximation. Econometrics 2019. www.mdpi.com/journal/econometrics (accessed Oct 6, 2020).

23 Sitas F, Newton R. Kaposi’s sarcoma in South Africa. J Natl Cancer Inst Monogr 2001; : 1–4.

24 Bray F, Colombet M, Mery L, et al., editors. Cancer Incidence in Five Continents, Vol. XI (electronic version). 2017 http://ci5.iarc.fr/CI5-XI/Default.aspx (accessed Jan 22, 2020).

25 Rue H, Martino S, Chopin N. Approximate {B}ayesian inference for latent {G}aussian models by using integrated nested {L}aplace approximations. J R Stat Soc Ser B (Statistical Methodol 2009; 71: 319–92.

26 Simbayi L, Zuma K, Zungu N, et al. South African National HIV Prevalence, Incidence and Behaviour and Communication Survey, 2017. Cape Town, 2019.

27 Rohner E, Bütikofer L, Schmidlin K, et al. Cervical cancer risk in women living with HIV across four continents: A multicohort study. Int J Cancer 2020; 146: 601–9.

28 National Cancer Registry. Cancer in South Africa 2016. 2020.

29 Akarolo-Anthony SN, Maso LD, Igbinoba F, Mbulaiteye SM, Adebamowo CA. Cancer burden among HIV-positive persons in Nigeria: Preliminary findings from the Nigerian AIDS-cancer match study. Infect Agent Cancer 2014; 9: 1.

30 Mbulaiteye SM, Katabira ET, Wabinga H, et al. Spectrum of cancers among HIV-infected persons in Africa: The Uganda AIDS-Cancer Registry Match Study. Int J Cancer 2006; 118: 985–90.

31 Shiels MS, Pfeiffer RM, Gail MH, et al. Cancer burden in the HIV-infected population in the United States. J Natl Cancer Inst 2011; 103: 753–62.

32 Franceschi S, Lise M, Clifford GM, et al. Changing patterns of cancer incidence in the early- and late-HAART periods: the Swiss HIV Cohort Study. Br J Cancer 2010; 103: 416–22.

33 Dryden-Peterson S, Medhin H, Kebabonye-Pusoentsi M, et al. Cancer incidence following expansion of HIV treatment in Botswana. PLoS One 2015; 10: 1–13.

34 Johnson LF, Rehle TM, Jooste S, Bekker LG. Rates of HIV testing and diagnosis in South Africa: Successes and challenges. AIDS 2015; 29: 1401–9.

35 Zaniewski E, Ostinelli CHD, Chammartin F, et al. Trends in CD4 and viral load testing 2005 to 2018: Multi-cohort study of people living with HIV in Southern Africa. medRxiv 2020; : 2020.03.09.20033423.

36 Kelly H, Weiss HA, Benavente Y, et al. Association of antiretroviral therapy with high-risk human papillomavirus, cervical intraepithelial neoplasia, and invasive cervical cancer in women living with HIV: a systematic review and meta-analysis. Lancet HIV 2018; 5: e45–58.

37 Menon S, Rossi R, Kariisa M, et al. Relationship between Highly Active Antiretroviral Therapy (HAART) and human papillomavirus type 16 (HPV 16) infection among women in Sub-Saharan Africa and public health implications: A systematic review. PLoS One 2019; 14: e0213086.

38 Bratcher LF, Sahasrabuddhe V V. The impact of antiretroviral therapy on HPV and cervical intraepithelial neoplasia: Current evidence and directions for future research. Infect. Agent. Cancer. 2010; 5: 8.

39 Horner MJ, Altekruse SF, Zou Z, Wideroff L, Katki HA, Stinchcomb DG. U.S. geographic distribution of prevaccine era cervical cancer screening, incidence, stage, and mortality. Cancer Epidemiol Biomarkers Prev 2011; 20: 591–9.

40 Lorenzo-Luaces Alvarez P, Guerra-Yi ME, Faes C, Galán Alvarez Y, Molenberghs G. Spatial analysis of breast and cervical cancer incidence in small geographical areas in Cuba, 1999-2003. Eur J Cancer Prev 2009; 18: 395–403.

41 Bingi D, Gidudu A, Okello D, Mwesigwa CL. Spatial Analysis of Cervical Cancer and Correlated Factors. J Remote Sens GIS 2018; 07. DOI:10.4172/2469-4134.1000223.

42 Thongsak N, Chitapanarux I, Suprasert P, et al. Spatial and Temporal Analyses of Cervical Cancer Patients in Upper Northern Thailand. Asian Pac J Cancer Prev 2016; 17: 5011–7.

43 Engels EA, Brock M V, Chen J, Hooker CM, Gillison M, Moore RD. Elevated incidence of lung cancer among HIV-infected individuals. J Clin Oncol 2006; 24: 1383–8.

44 Zhao J, Virani S, Sriplung H. Spatiotemporal mapping of cervical cancer incidence highlights need for targeted prevention in Songkhla province, Thailand. Health Policy Plan 2017; 32: 430–6.

45 Zhan FB, Lin Y. Racial/Ethnic, socioeconomic, and geographic disparities of cervical cancer advanced-stage diagnosis in Texas. Womens Health Issues 2014; 24: 519–27.

46 Fusheini A, Eyles J. Achieving universal health coverage in South Africa through a district health system approach: conflicting ideologies of health care provision. BMC Health Serv Res 2016; 16: 1–11.

47 Ataguba JEO, McIntyre D. Who benefits from health services in South Africa? Heal Econ Policy Law 2013; 8: 21–46.

48 Spees LP, Brewster WR, Varia MA, et al. Examining Urban and Rural Differences in How Distance to Care Influences the Initiation and Completion of Treatment among Insured Cervical Cancer Patients. Cancer Epidemiol Biomarkers Prev 2019; 28: 882–9.

49 Matenge TG, Mash B. Barriers to accessing cervical cancer screening among HIV positive women in Kgatleng district, Botswana: A qualitative study. PLoS One 2018; 13: e0205425.

50 Akinyemiju TF, McDonald JA, Lantz PM. Health care access dimensions and cervical cancer screening in South Africa: analysis of the world health survey. BMC Public Health 2015; 15: 382.

