## Supplementary material for "Spatiotemporal modelling and mapping of cervical cancer incidence among HIV positive women in South Africa: A nationwide study": Table S1.1

### Online Supplement: Spatiotemporal mapping of cervical cancer incidence among women living with HIV in South Africa: A nationwide study

Tafadzwa Dhokotera[1], Julien Riou[1], Lina Bartels[1], Eliane Rohner[1], Frederique Chammartin[1], Elvira Sign[2,4], Victor Olago[2,4], Mazvita Sengavi[2,4], Matthias Egger[1], Julia Bohlius[1], and Garyfallos Konstantinoudis*[1,5]

[1]*Institute of Social and Preventive Medicine (ISPM), University of Bern, Bern*

[2]*Centre for Infectious Disease Epidemiology and Research, University of Cape Town, South Africa*

[3]*Centre for Infectious Disease Epidemiology and Research, School of Public Health and Family Medicine, University of Cape Town, South Africa*

[4]*Division of Epidemiology and Biostatistics, School of Public Health, University of the Witwatersrand, Johannesburg, South Africa*

[5]*Epidemiology and Biostatistics department, School of Public Health, Imperial College London, United Kingdom*

### 1 Text

#### 1.1 Thembisa model disaggregation

The Thembisa model (among other information) provides estimates of HIV prevalent cases and people diagnosed with HIV per province, year and age during 2004 and 2014. For our analysis we used people diagnoses with HIV as the denominator. To disaggregate the provincial HIV counts to the municipality unit we calculated weights based on information provided by the National Health Laboratory Service (NHLS). We focused on individuals with $\geq 2$ tests, and we selected as date of HIV diagnosis the date of the first recorded test/laboratory result. We assumed that the municipality where the first test was performed is the same as the municipality of residence of the HIV case.

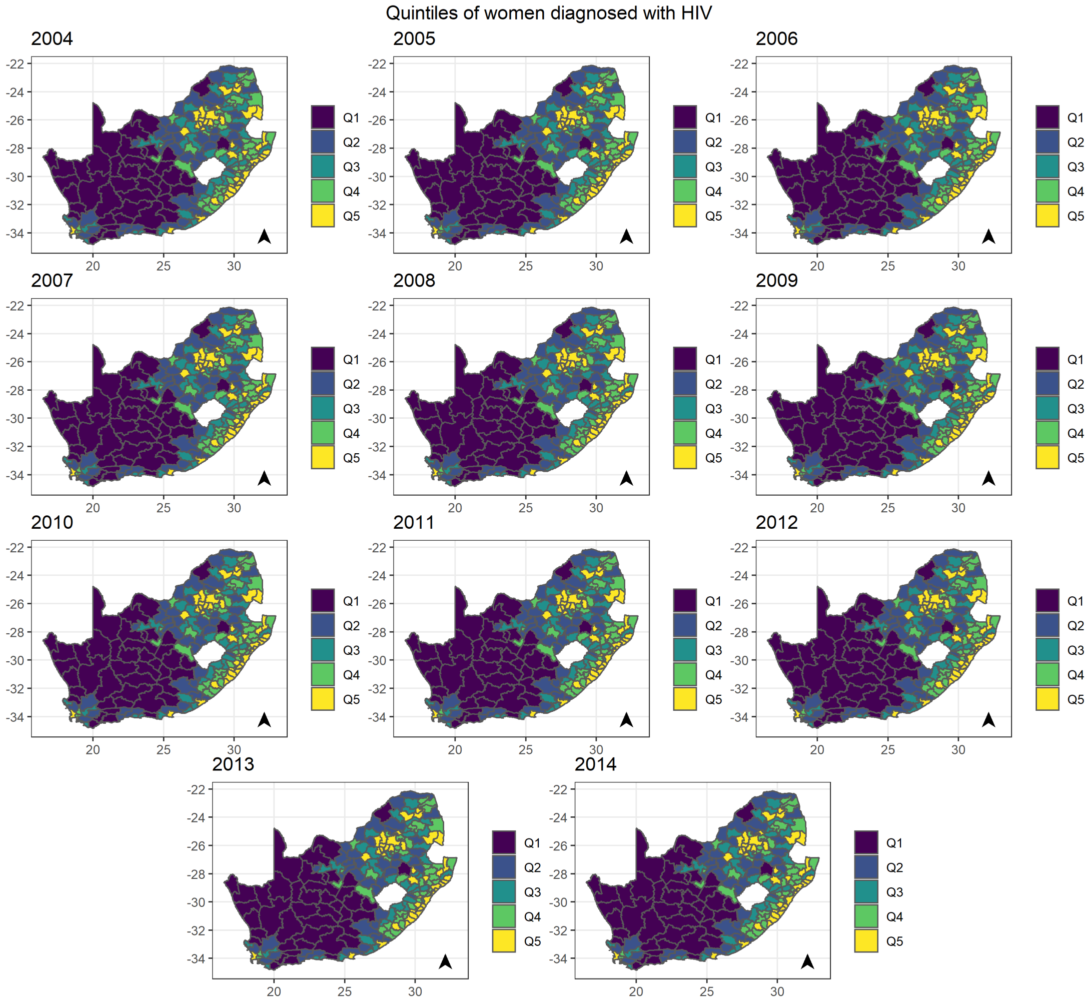

Figure 1. The quintiles of the population density per municipality of women diagnosed with HIV as resulted from disaggregating the Thembisa model using weights calculated from the National Health Laboratory Service dataset.

Let $P_{ijt}$ be the number of women diagnosed with HIV (based on NHLS) in the $i$-th municipality where $i = 1, \ldots, n$, $j$-th province, with $j = 1, \ldots, J$ and at time $t$, $t = 1, \ldots, T$. We calculated province-specific municipality weights as $w_{ijt} = P_{ijt}/P_{\cdot jt}$ where $P_{\cdot jt} = \sum_{i \sim j} P_{ijt}$ where $i \sim j$ denotes the municipalities that belong to the $j$-th province at time $t$. To dissagregate the Thembisa model provincial counts $\tilde{P}_{\cdot jt}$ at time $t$, we multiplied them with the weights $w_{ijt}$, i.e. $\tilde{P}_{it} = w_{ijt} \cdot \tilde{P}_{\cdot jt}$.
To get the age dimension, we assumed that $w_{ijt}$ is constant over the different $k$ age groups considered (0-4, 5-9, ..., >80) and retrieved $\tilde{P}_{itk}$. The output of the procedure is given on Figure 1 for all ages.

#### 1.2 Model description

Let $A$ be an observation window divided in spatial units $A_1, A_2 \dots A_n$ (municipalities in South Africa). Let $Y_{itk}$ be the counts of cervical cancer cases in the $i$-th municipality, $t$-th year and $k$-th age group. A general model formulation would be:

$$
\begin{aligned}
Y_{itk}|\lambda_{itk},\tilde{P}_{itk} &\sim \text{Poisson}(\lambda_{itk}\tilde{P}_{itk}) \\
\log(\lambda_{itk}) &= \beta_0 + \eta_k + w_t + \phi_i + \delta_{it} \\
\eta_k &\sim \text{RW1}(\sigma_1^2) \\
\phi_i &\sim \text{BYM2}(\boldsymbol{W}, \sigma_2^2, \rho) \\
w_t &\sim \text{RW1}(\sigma_3^2) \\
\delta_{it} &\sim \mathcal{N}(0, \sigma_4^2) \\
\beta_0 &\sim \mathcal{N}(0, \infty) \\
\sigma_1^2, \sigma_2^2, \sigma_3^2, \rho &\sim \text{PCpriors}
\end{aligned}
$$

where $\beta_0$ is an intercept term, $\eta$ the age group random effect defined as a random walk of order 1 (RW1), $\phi$ the spatial random effect [Besag et al., 1991, Simpson et al., 2017], $w_t$ a temporally strucutred random effect (RW1), $\delta$ the spacetime interaction (we considered type I) of the spatial and temporal components. The type I interaction refers to unstructured overdispersion in time and space Knorr-Held [2000]. The hyperparameters $\sigma_1^2, \sigma_2^2, \sigma_3^2$, $\sigma_4^2$ are variances, $\rho$ is the mixing parameter of the spatial field $\phi_i$ and $\mathbf{W}$ the neighborhood matrix.

The priors for all the variance hyperparameters where set based on $\Pr(\sigma_i < 1) = 0.01$, for $i = 1, 2, 3, 4$ reflecting that is unlikely to have a risk $\exp(1) \approx 2.72$ times higher than the temporal, age specific, spatial average or spatiotemporal average. The mixing parameter $\rho$ of the spatial field was selected based on $\Pr(\rho > 1) = 0.50$, reflecting our lack of knowledge whether the unstructured or spatially structured random effect should dominate the field. For more information about the PCpriors see Simpson et al. [2017].

#### 1.3 Correction II

Let $i = 1, \ldots, n$ be the number of municipalities, $j = 1, \ldots, J$ the provinces in South Africa and $t = 1, \ldots, T$ for the years. Let $C_{ijtl}$ stand for the number of Kaposi sarcoma (KS) cases residing in the $i$ municipality, with their lab report sent to the $j$-th province in the $t$-th year and $l = 1$ to denote the linked cases (with the NHLS after performing the linkage) and 0 the unlinked.

For $l = 0$, the cases are linked with the NHLS thus the municipality of residence is known, and thus $C_{it1}$ is known. However, for the unlinked cases we only have the province where the cancer test was sent. Thus we do not have the $C_{it0}$, but we have $C_{jt0}$. To calculate $C_{it0}$, we calculated weights defined as: $w_{ijt} = C_{ijt1}/C_{jt1}$. The interpretation of these weights is: the proportion of tests (for cancer diagnosis) of the $i$-th municipality that are sent to the $j$-th province to be examined (at time $t$ among the linked cancer cases). To approximate $C_{it0}$, we define $\tilde{C}_{it0} \approx \sum_j w_{ijt} = C_{jt0}$. Thus the correction factor is:

$$b_{it} = \frac{C_{it1}}{C_{it1} + \tilde{C}_{it0}}$$

We additionally excluded the KS cases not linked and treated in the private sector. Let $K_{it0}$ be the KS cases not linked with NHLS and treated in the private sector, we can then write:

$$b_{it} = \frac{C_{it1} + K_{it0}}{C_{it1} + \tilde{C}_{it0}}$$

The output of the above procedure is shown in Figure 2. We note, that we aggregate in time to avoid having a lot of zeros in the data, making it hard to apply the correction on the model-based incidence output.

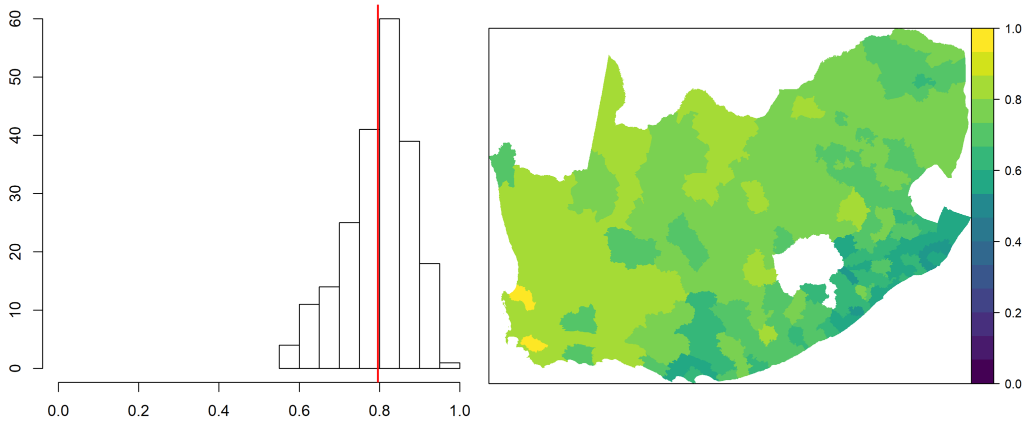

Figure 2. The correction II factor in space (right panel) and its histogram (left panel).

Table S1: Deviance information criterion (DIC), Watanabe-Akaike information criterion (WAIC) and mean logarithmic score (CPO) for the different models considered. For the notation refer to Text 1.2.

| models | DIC | WAIC | CPO |
|---|---|---|---|
| $\beta_0$ | 56694.15 | 56700.84 | 1.09 |
| $\beta_0 + \eta_k$ | 35668.98 | 35685.81 | 0.69 |
| $\beta_0 + w_t$ | 56419.44 | 56480.29 | 1.08 |
| $\beta_0 + \phi_i$ | 52535.31 | 52699.83 | 1.01 |
| $\beta_0 + \eta_k + w_t$ | 34110.17 | 34133.37 | 0.66 |
| $\beta_0 + \eta_k + \phi_i$ | 31559.09 | 31606.54 | 0.61 |
| $\beta_0 + w_t + \phi_i$ | 52276.15 | 52495.72 | 1.01 |
| $\beta_0 + \eta_k + w_t + \phi_i$ | 30061.23 | 30093.06 | 0.58 |
| $\beta_0 + w_t + \phi_i + \delta_{it}$ | 51853.77 | 53005.64 | 1.02 |
| $\beta_0 + \eta_k + w_t + \phi_i + \delta_{it}$ | 29625.55 | 29726.33 | 0.57 |

$\beta_0$ is an intercept term, $\eta_k$ the age effect, $w_t$ the temporal effect, $\phi_i$ the spatial effect and $\delta_{it}$ the spatiotemporal effect.

Table S2: Annual median and 95% Credibility intervals (CrI) for the incidence rate of cervical cancers among HIV positive women per 100,000 person years in South Africa for the different corrections considered.

| | No correction | | Correction I | | Correction II | | Full Correction | |
|---|---|---|---|---|---|---|---|---|
| year | Median | 95% CrI | Median | 95% CrI | Median | 95% CrI | Median | 95% CrI |
| 2004 | 306 | (169, 555) | 306 | (163, 573) | 310 | (164, 589) | 312 | (160, 609) |
| 2005 | 386 | (221, 675) | 378 | (211, 680) | 398 | (224, 718) | 394 | (217, 727) |
| 2006 | 417 | (243, 718) | 407 | (231, 720) | 436 | (249, 771) | 430 | (241, 775) |
| 2007 | 341 | (198, 588) | 336 | (190, 593) | 357 | (203, 630) | 354 | (198, 636) |
| 2008 | 319 | (187, 543) | 318 | (183, 553) | 340 | (197, 590) | 341 | (195, 603) |
| 2009 | 294 | (174, 498) | 297 | (172, 514) | 314 | (183, 542) | 319 | (183, 559) |
| 2010 | 269 | (160, 451) | 272 | (159, 467) | 289 | (170, 494) | 294 | (171, 510) |
| 2011 | 231 | (139, 386) | 237 | (140, 403) | 249 | (149, 422) | 256 | (151, 440) |
| 2012 | 203 | (122, 338) | 209 | (123, 353) | 218 | (130, 367) | 224 | (132, 383) |
| 2013 | 179 | (108, 296) | 187 | (111, 316) | 193 | (116, 324) | 202 | (120, 344) |
| 2014 | 160 | (96, 265) | 179 | (106, 303) | 172 | (103, 290) | 191 | (113, 326) |
| Median | 294 | (174, 498) | 297 | (163, 573) | 310 | (170, 494) | 312 | (171, 510) |

Table S3: Results of the model with spatial, temporal and spatiotemporal interaction and deprivation and urbanicity for the different Thembisa denominators.

| | Univariable | | Multivariable | |
|---|---|---|---|---|
| | median | 95% CrI | median | 95% CrI |
| $1/\sigma_1^{2*}$ | 32.36 | (10.93, 83.24) | 33.05 | (11.88, 83.25) |
| $1/\sigma_2^{2*}$ | 3.53 | (2.52, 4.84) | 4.07 | (2.75, 5.94) |
| $\phi^{2*}$ | 0.62 | (0.36, 0.84) | 0.77 | (0.45, 0.95) |
| $1/\sigma_3^{2*}$ | 1.28 | (0.64, 2.41) | 1.28 | (0.64, 2.43) |
| $1/\sigma_4^{2*}$ | 16.22 | (12.82, 20.69) | 16.05 | (12.69, 20.41) |

CrI: Credibility intervals

$1/\sigma_1^2$ is the precision of the random walk of order 1 (RW1) of the age effect, $1/\sigma_2^2$ of the spatial field, $1/\sigma_3^2$ of the temporal effect, and $1/\sigma_4^2$ of the spatiotemporal interaction.

* The hyperparameters refer to the distribution of the logged random effects.

Figure S1: Flowchart for the exclusion criteria used to calculate weights using data from the National Health Laboratory Service (NHLS) to disaggregate the Thembisa provincial estimates.

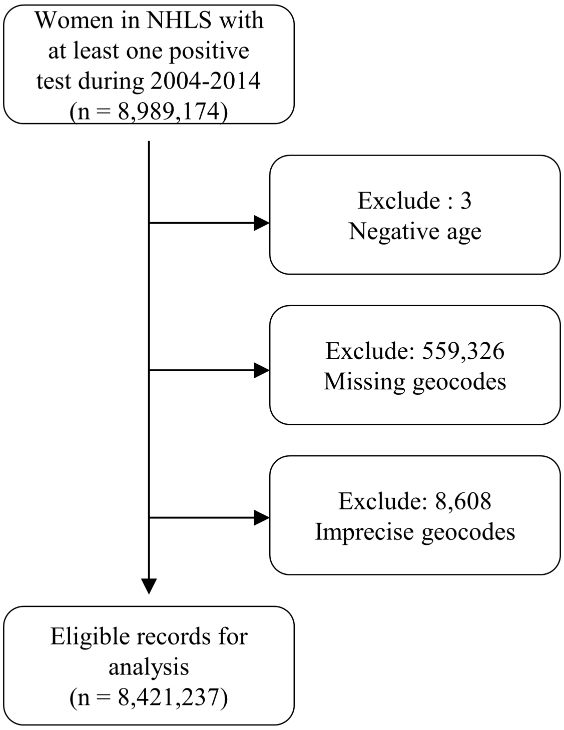

Figure S2: The spatial variation of urbanicity (urban/rural) in South Africa.

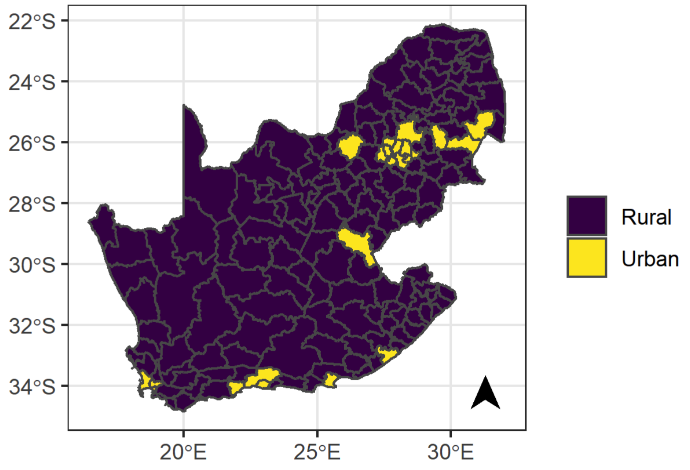

Figure S3: Provinces in South Africa in 2016.

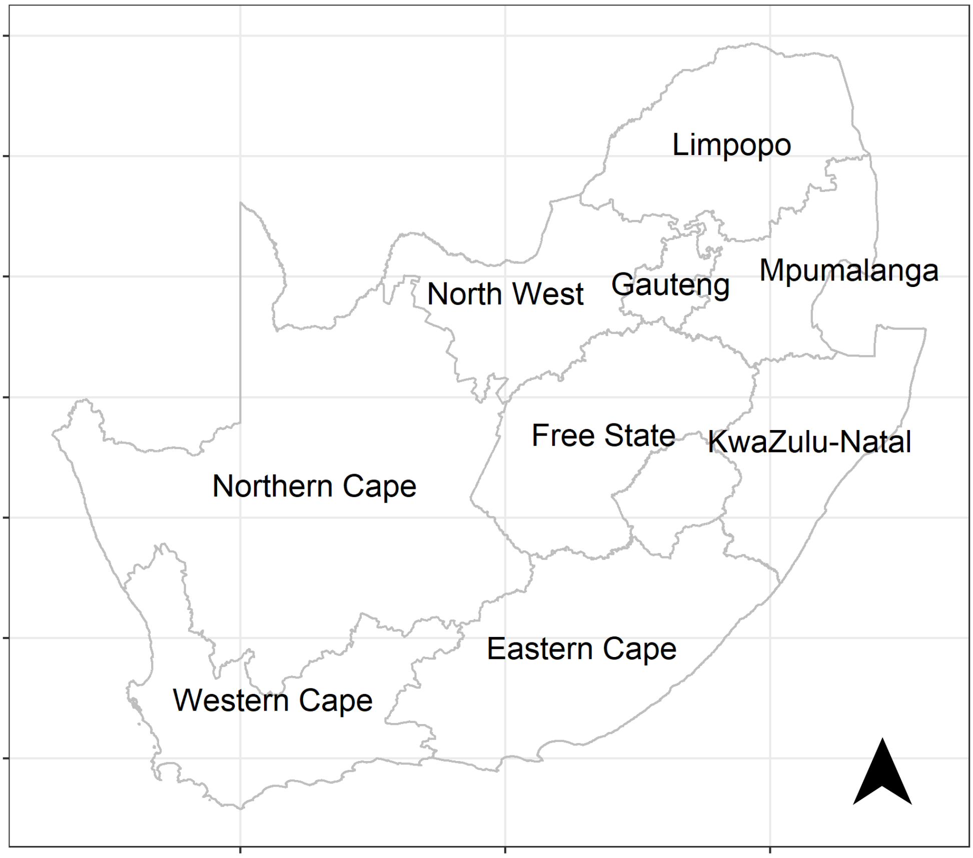

Figure S4: Posterior probability that the spatiotemporal relative risk (relative to the national average over time) is higher than 1 of cervical cancers among women living with HIV in South Africa, using the no correction model without any covariates.

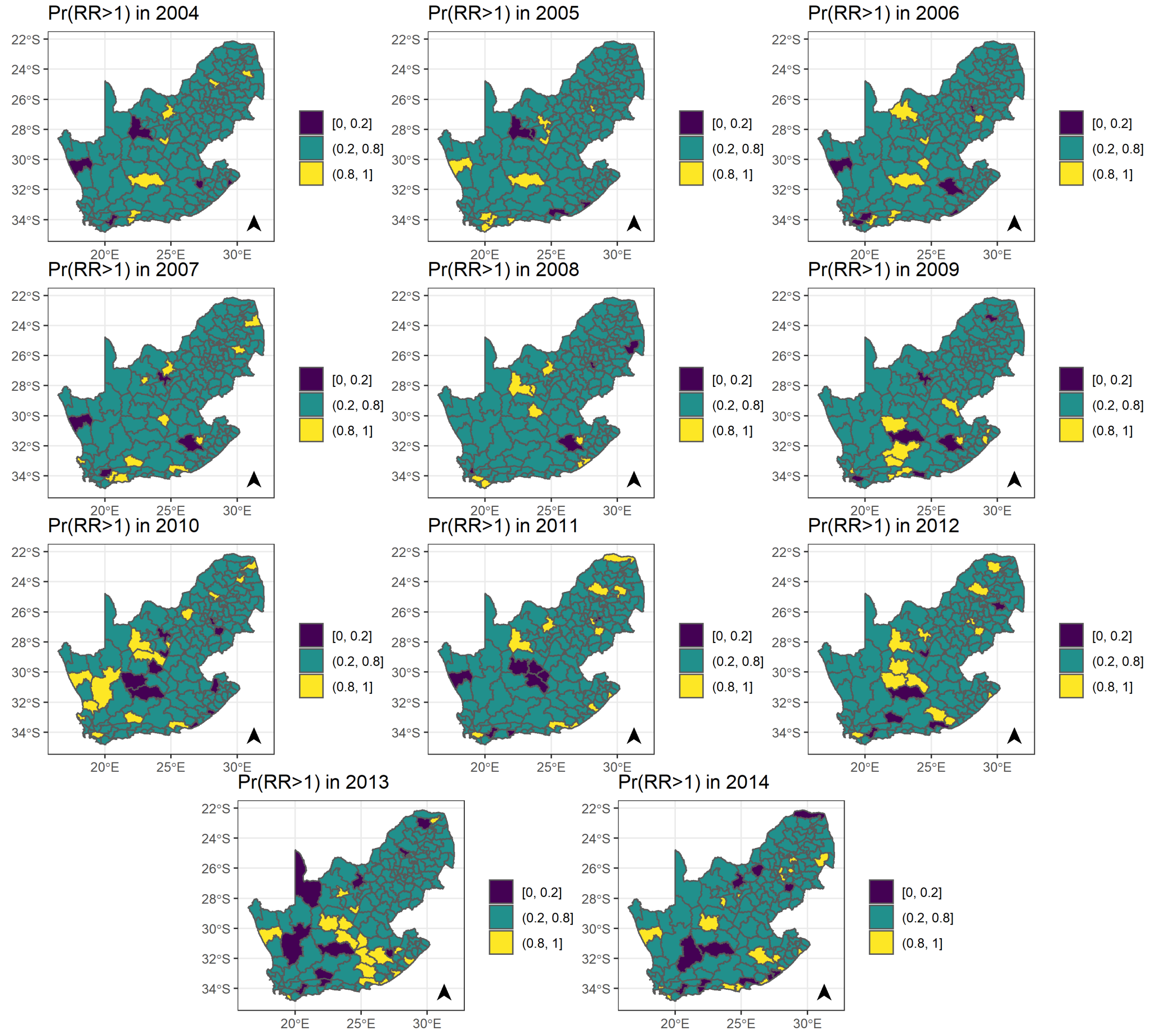

Figure S5: Posterior probability that the spatiotemporal relative risk (relative to the national average over time) is higher than 1 of cervical cancers among women living with HIV in South Africa, using the no correction model adjusted for the selected covariates.

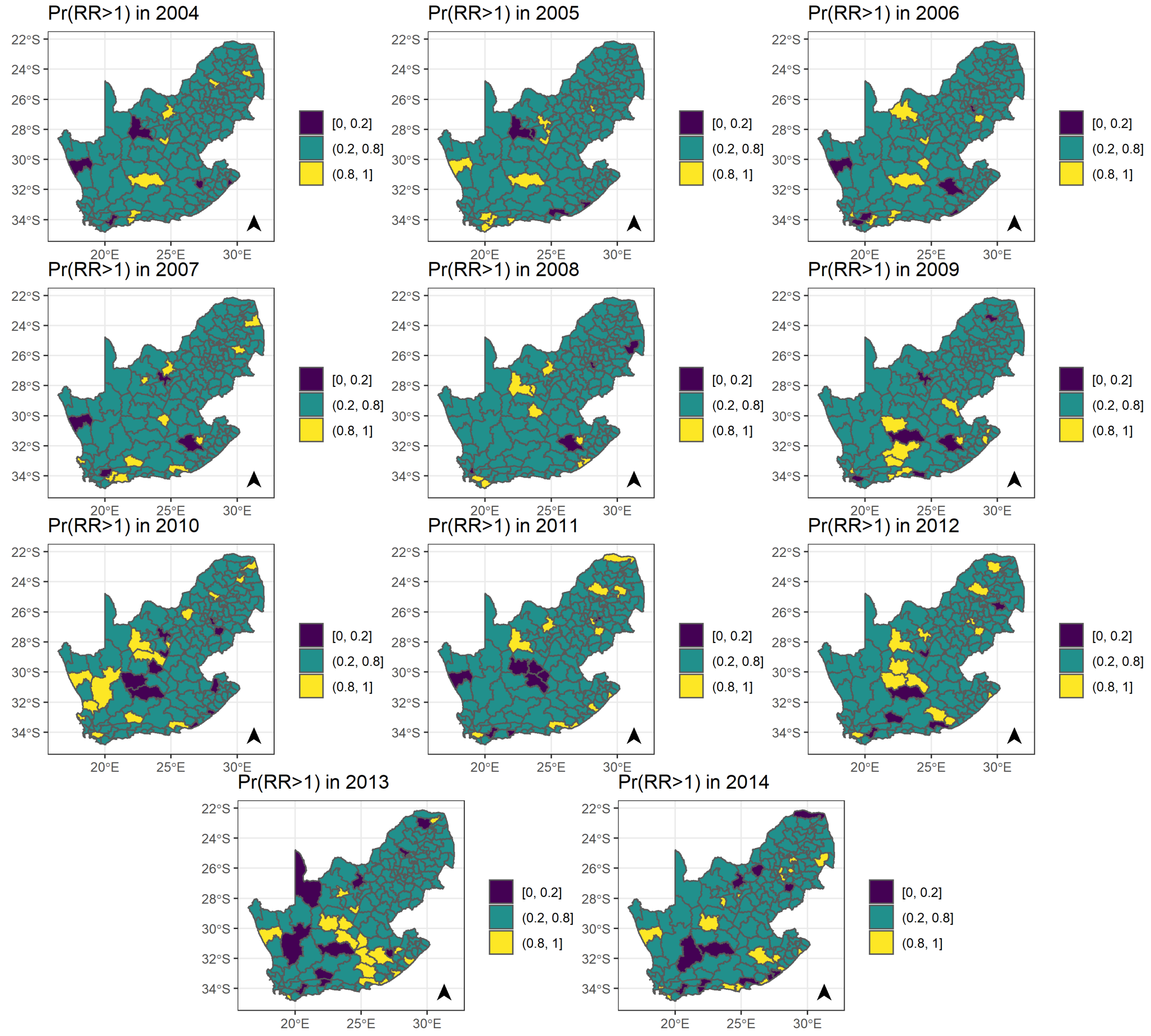

### References

Julian Besag, Jeremy York, and Annie Mollié. Bayesian image restoration, with two applications in spatial statistics. *Annals of the institute of statistical mathematics*, 43(1):1–20, 1991.

Leonhard Knorr-Held. Bayesian modelling of inseparable space-time variation in disease risk. *Statistics in medicine*, 19(17-18):2555–2567, 2000.

Daniel Simpson, Håvard Rue, Andrea Riebler, Thiago G Martins, Sigrunn H Sørbye, et al. Penalising model component complexity: A principled, practical approach to constructing priors. *Statistical science*, 32(1):1–28, 2017.
